# Diabetes is associated with increased risk for in-hospital mortality in patients with COVID-19: a systematic review and meta-analysis comprising 18,506 patients

**DOI:** 10.1101/2020.05.26.20113811

**Authors:** Leonidas Palaiodimos, Natalia Chamorro-Pareja, Dimitrios Karamanis, Weijia Li, Phaedon D. Zavras, Priyanka Mathias, Damianos G. Kokkinidis

**Author notes:** Corresponding author: Leonidas Palaiodimos, MD MSc, Division of Hospital Medicine, Montefiore Medical Center, Albert Einstein College of Medicine, 111 East 210^th^ street, Bronx, NY 10467, NW building, 8^th^ floor.

## Abstract

**Background:** Infectious diseases are more frequent and can be associated with worse outcomes in patients with diabetes. Our aim was to systematically review and synthesize with a meta-analysis the available observational studies reporting the effect of diabetes in mortality among hospitalized patients with COVID-19.

**Methods:** Medline, Embase, Google Scholar, and medRxiv databases were reviewed. A random-effect model meta-analysis was used and I-square was utilized to assess the heterogeneity. In-hospital mortality was defined as the endpoint. Sensitivity, subgroup, and meta-regression analyses were performed.

**Results:** 18,506 patients were included in this meta-analysis (3,713 diabetics and 14,793 non-diabetics). Patients with diabetes were associated with a higher risk of death compared to patients without diabetes (OR: 1.65; 95% CI: 1.35-1.96; I^2^ 77.4%). The heterogeneity was high. A study level meta-regression analysis was performed for all the important covariates and no significant interactions were found between the covariates and the outcome of mortality.

**Conclusion:** This meta-analysis shows that that the likelihood of death is 65% higher in diabetic hospitalized patients with COVID-19 compared to non-diabetics. Further studies are needed to assess whether this association is independent or not, as well as to investigate to role of glucose control prior or during the disease.

## Introduction

Diabetes Mellitus is one of the leading causes of morbidity and mortality in the world and is mainly associated with significant cardiovascular and renal complications. The estimated global prevalence was 9.3% in 2019 with an increasing trend (1, 2). In the United States alone, more than 34 million adults had known or undiagnosed diabetes in 2018 (3). In 2017, diabetes was listed as the underlying or contributing cause of death on 270,702 death certificates that corresponds to a crude rate of 83.1 per 100,000 persons (3).

Infectious diseases are more frequent and can be associated with worse outcomes in patients with diabetes (4). Therefore, it is not surprising that diabetes has been considered a possible risk factor or a predictor for worse outcomes in patients with Coronavirus Disease 2019 (COVID-19) (5-7). COVID-19 rapidly reached the level of pandemic and has caused more than 300 thousand deaths worldwide within a few months despite unprecedented mitigation measures (8). The strength of the association between the two pandemics, diabetes and COVID-19, has been investigated in observational cohorts around the world.

We aimed to systematically review and synthesize with a meta-analysis the available observational studies reporting the effect of diabetes in mortality among hospitalized patients with COVID-19.

## Methods

This study was performed according to the Preferred Reporting Items for Systematic Reviews and Meta-Analyses (PRISMA) (9).

### Literature search

We conducted a systematic literature search of the Medline, Embase, Google Scholar, and medRxiv (the preprint server for health sciences) up to May 10, 2020 for observational studies providing any kind of association between diabetes and mortality in hospitalized patients with COVID-19. Two investigators (LP, DGK) independently searched for eligible studies. In cases where there was a disagreement regarding the eligibility of a study, a third investigator (FZ) was involved in order consensus to be reached. The reference list of pertinent reviews and observational studies was also manually searched for further potentially eligible studies. A combination of the following keywords was used to perform our searches: “COVID-19”, “SARS-CoV-2”, “novel coronavirus”, “risk factor”, “mortality”, and “death”. The pre-specified inclusion criteria were: (i) studies included adult patients hospitalized for COVID-19, (ii) studies provided any kind of association between diabetes and mortality in the aforementioned population. The prespecified exclusion criteria were: (i) certainly or possibly duplicated or overlapping patient populations and (ii) studies included prespecified patient populations based on a specific diagnosis (e.g. only hypertensives or only patients with cancer). In the cases of duplicated or overlapping populations, the studies with the larger samples were included.

### Data extraction and outcomes

Data extraction was performed based on a pre-defined data extraction form by two independent investigators (NCP, WL) blinded to each other. The pre-specified outcome was in-hospital mortality.

### Risk of bias assessment

Two independent reviewers (NCP, PM) assessed the risk of bias of the included studies with Quality in Prognosis Studies tool (QUIPS) (10). Studies were assessed as having low, moderate, serious or critical risk of bias for the following domains: study participation, study attrition, prognostic factor measurement, confounding measurement and account, outcome measurement, analysis and reporting.

### Statistical analysis

We estimated the odds ratios (ORs) and their respective 95% confidence intervals (CI) for all the individual studies. When neither OR nor event rates were provided, we used the unadjusted hazard ratios and converted them to ORs given the short follow-up period. For studies that provided both adjusted and unadjusted ORs, we used the unadjusted effect estimate. We performed a meta-analysis using the random effects model according to the method of Der Simonian and Laird. Heterogeneity among trials for each outcome was assessed with the I-squared test. Values < 25% indicated low, 25 to 70% moderate, and > 70% high heterogeneity. Egger test and funnel plots were used to assess for publication bias. Subgroup and sensitivity analyses were performed based on the location where the studies were conducted and the mean/median age. A meta-regression analysis was performed for important covariates in order to address high heterogeneity among included studies. Statistical significance level was set at 0.05 with CI calculated at the 95% level. Stata 14.1 (Stata Corp., College Station, Texas) was used for statistical analysis.

## Results

Out of 1,721 studies screened from the literature and on-line sources, fourteen observational studies (twelve retrospective and two prospective) met the prespecified criteria for inclusion in the analysis **(Figure 1)** (11-24). The characteristics of these studies are summarized in **Table 1**. Overall, all the studies were found to have low risk of bias **(Figure 2)**. Five studies were conducted in Asia, five in the United States and four in Europe. The total number of patients included in the final dataset was 18,506 patients: 3,713 diabetics and 14,793 non-diabetics. The mean or median age was above 60 years in twelve studies. 43% (7,967) of the population was women. Among studies which reported their results in event rates, the overall frequency of death events in diabetics was 41.1% (991 out of 2,413) compared to 17.6% (2,113 out of 12,012) in the non-diabetic group (the numbers of death events in the groups of interest were not provided in three studies; instead, odds or hazard ratios were provided). In our meta-analysis of 14 studies, we found that patients with diabetes were associated with a higher risk of death compared to patients without diabetes, but with significant heterogeneity (OR: 1.65; 95% CI: 1.35-1.96; I^2^ 77.4%; **Figure 3)**.

**Figure 1.**
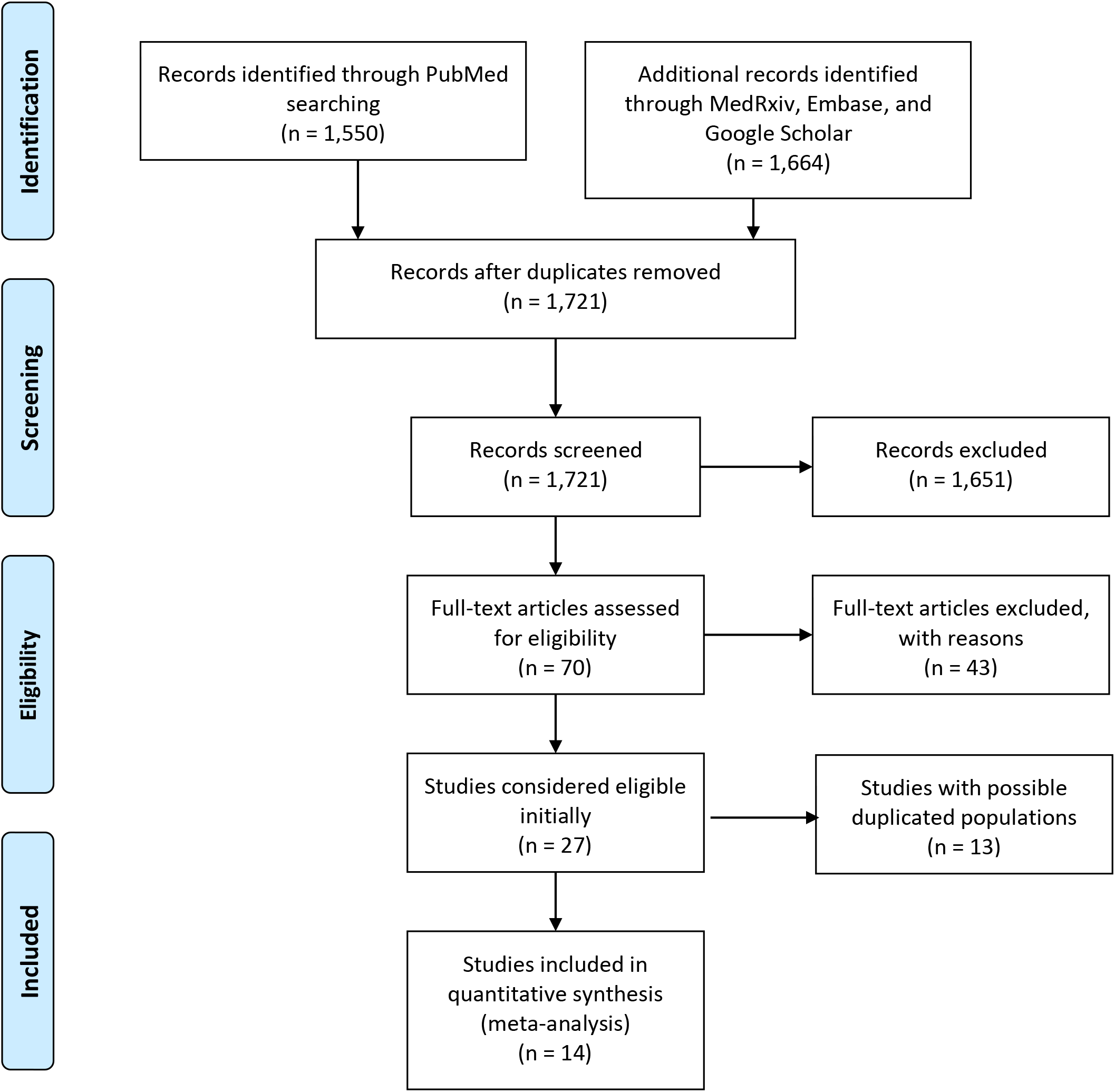
PRISMA 2009 flow diagram

**Table 1:** Characteristics of the included studies.

| Study | Country | Region | Institution | Study design | First patient | Last patient | Number of included patients |
| --- | --- | --- | --- | --- | --- | --- | --- |
| **Guan** | China | 31 regions (including Wuhan) | Multicenter | Retrospective | December 25 | January 31 | 1,590 |
| **Han** | China | Wuhan | Tongji Hospital | Retrospective | February 2 | February 15 | 306 |
| **Zeng** | China | Xi'an and Wuhan | Multicenter (not Tongji hospital) | Retrospective | February 5 | March 20 | 97 |
| **Nikpouraghdam** | Iran | Tehran | Baqiyatallah Hospital | Retrospective | February 19 | April 15 | 2,964 |
| **Javanian** | Iran | Babol | Babol University of Medical Sciences | Retrospective | February 25 | March 12 | 100 |
| **Rossi** | Italy | Reggio Emilia | Multicenter | Prospective | February 27 | April 2 | 1,075<sup>a</sup> |
| **Borobia** | Spain | Madrid | La Paz University Hospital | Retrospective | February 25 | April 19 | 2,226 |
| **Perez Guzman** | United Kingdom | London | Imperial College Healthcare NHS Trust | Retrospective | February 25 | April 5 | 520 |
| **Tomlins** | United Kingdom | Bristol | North Bristol NHS Trust | Retrospective | March 1 | March 30 | 95 |
| **Levy** | United States | New York | Northwell Health | Retrospective | March 1 | April 12 | 5,233 |
| **Wang** | United States | New York | Mount Sinai Health System | Retrospective | March 7 | April 15 | 3,273 |
| **Cummings** | United States | New York | New York-Presbyterian | Prospective | March 2 | April 1 | 257 |
| **Palaiodimos** | United States | New York | Montefiore Medical Center | Retrospective | March 9 | March 22 | 200 |
| **Bode** | United States | 10 states (not New York) | Multicenter | Retrospective | March 1 | April 6 | 570<sup>b</sup> |

Notes: a. Only the subset of patients who were hospitalized were included, b. Only the subset of patients who had been discharged or died were included

**Figure 2.**
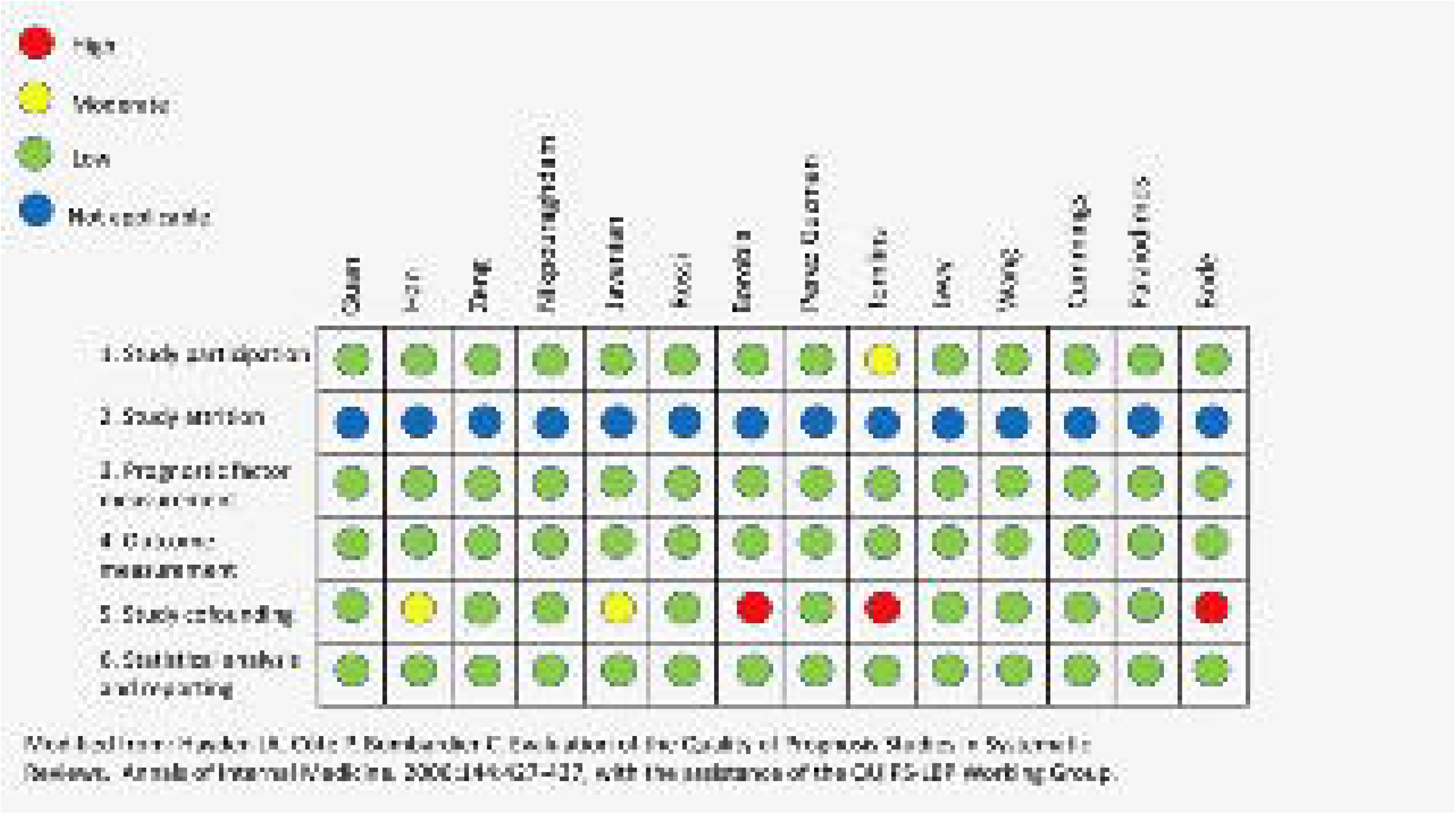
Risk of bias assessment based on the Quality in Prognosis Studies (QUIPS) tool

**Figure 3.**
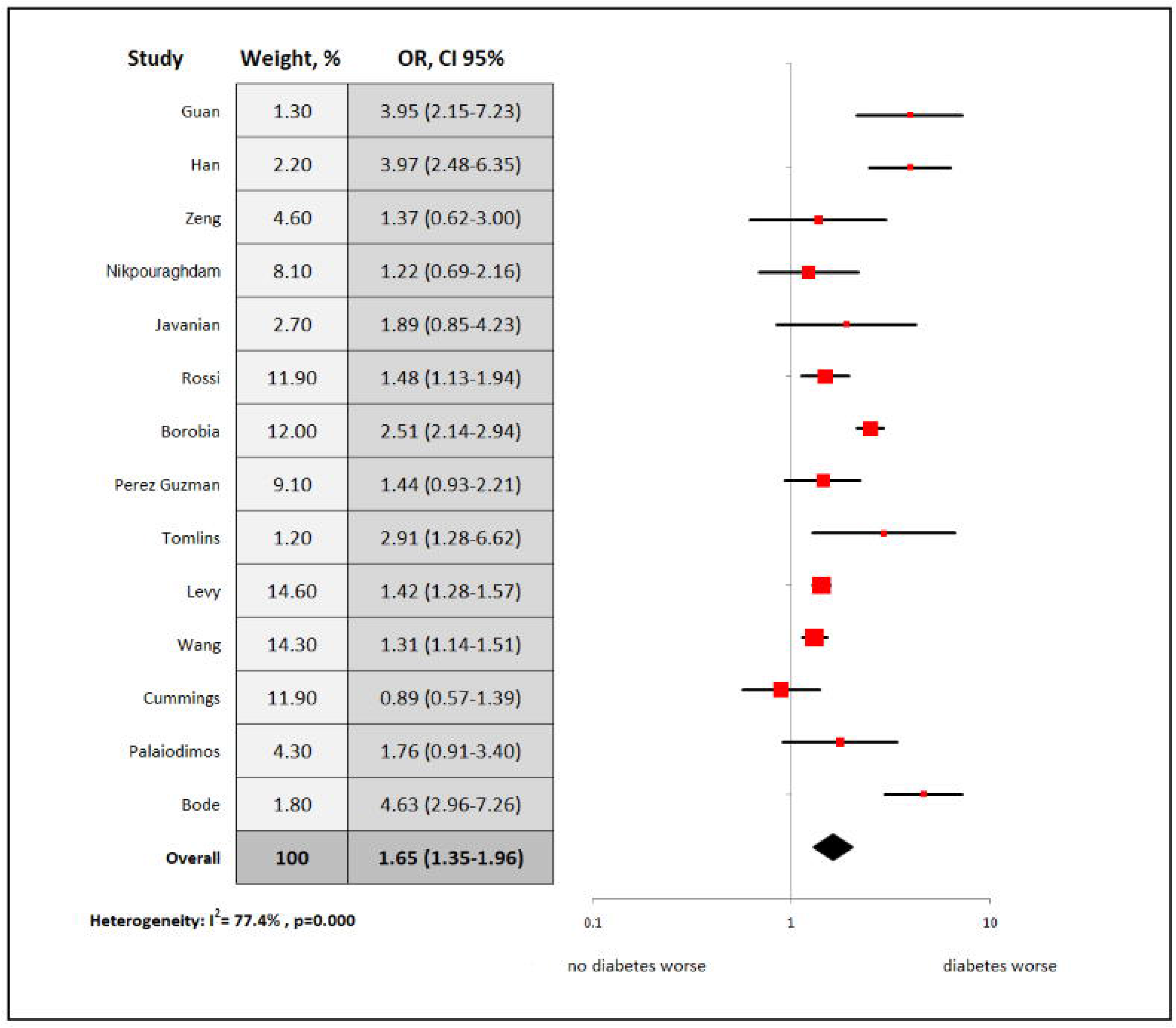
Overall analysis: diabetes vs. no diabetes for in-hospital mortality

### Sensitivity and subgroup analyses

Sensitivity analysis were conducted for studies that were performed in the United States (N = 5), Asia (5), and Europe-United States (N=9). Similarly, to the overall analysis, our sensitivity analysis for studies conducted in the United States revealed a higher chance of death among diabetic patients compared to non-diabetes group (OR: 1.34; 95% CI: 1.04-1.85; I^2^ 73.7%). We found a similar association for studies conducted in Asia (OR: 2.12; 95% CI: 1.09-3.16; I^2^ 61.1%) and among studies conducted in the United States or Europe (OR: 1.60; 95% CI: 1.27-1.93; I^2^ 82.8%, **Figure 4)**. Subgroup analysis of the studies that had a mean or median age less than 60 years (N=2, both from Asia) did not show a significant difference in mortality between diabetics and non-diabetics (OR: 2.3; 95% CI: 0.01-4.92; I^2^ 75.5%, **Figure 4)** but was probably limited by the small sample given the wide confidence intervals.

**Figure 4.**
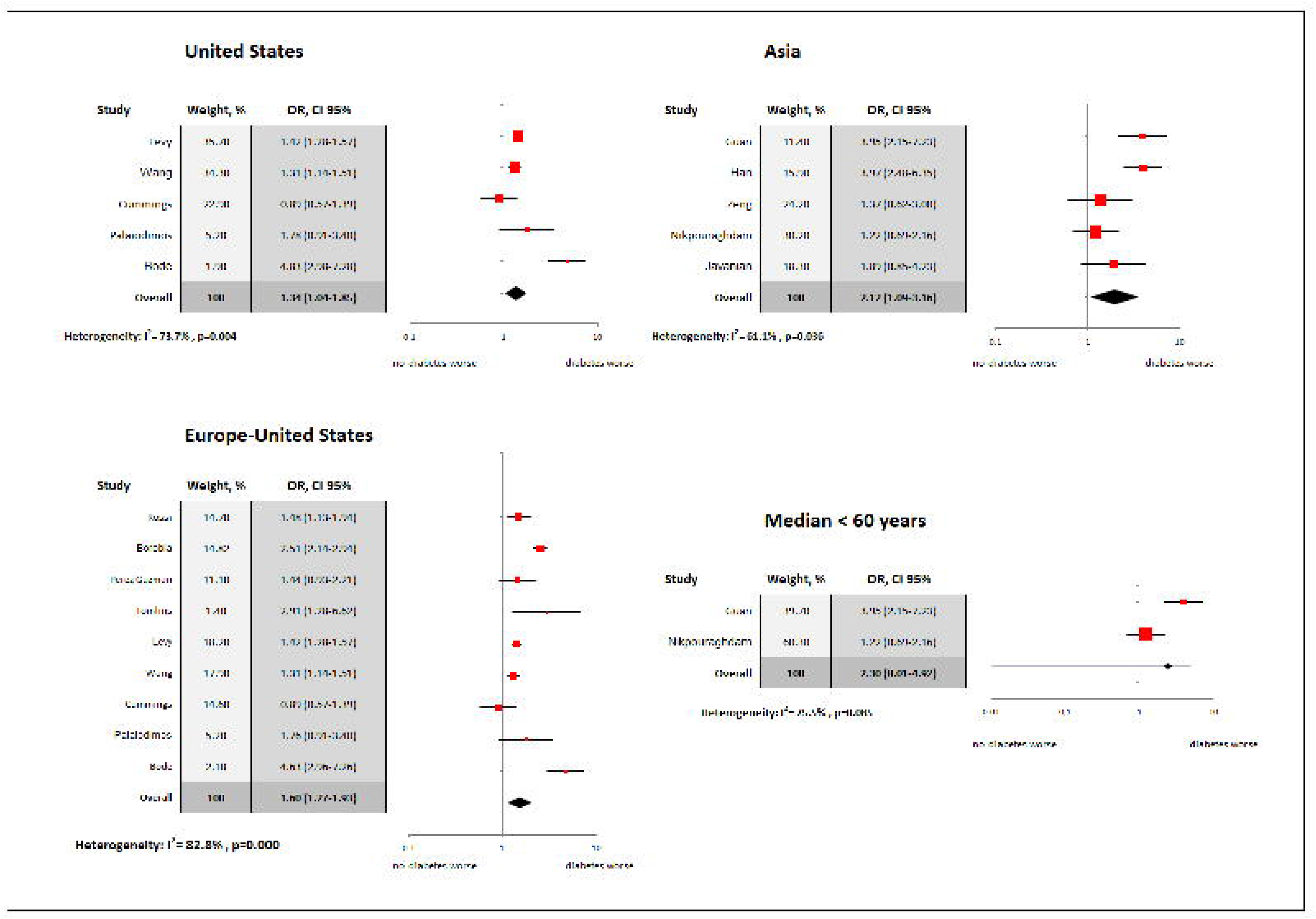
Sensitivity and subgroup analyses based on the region of the study origin and the mean/median age of the study population: diabetes vs. no diabetes for in-hospital mortality

### Assessment for Publication Bias

Assessment for publication bias was performed with two different ways. First, an Egger test was conducted that was not suggestive of publication bias (P = 0.255). Second, we performed a visual assessment of the results of the funnel plot which was not indicative of publication bias, as well **(Figure 5)**.

**Figure 5.**
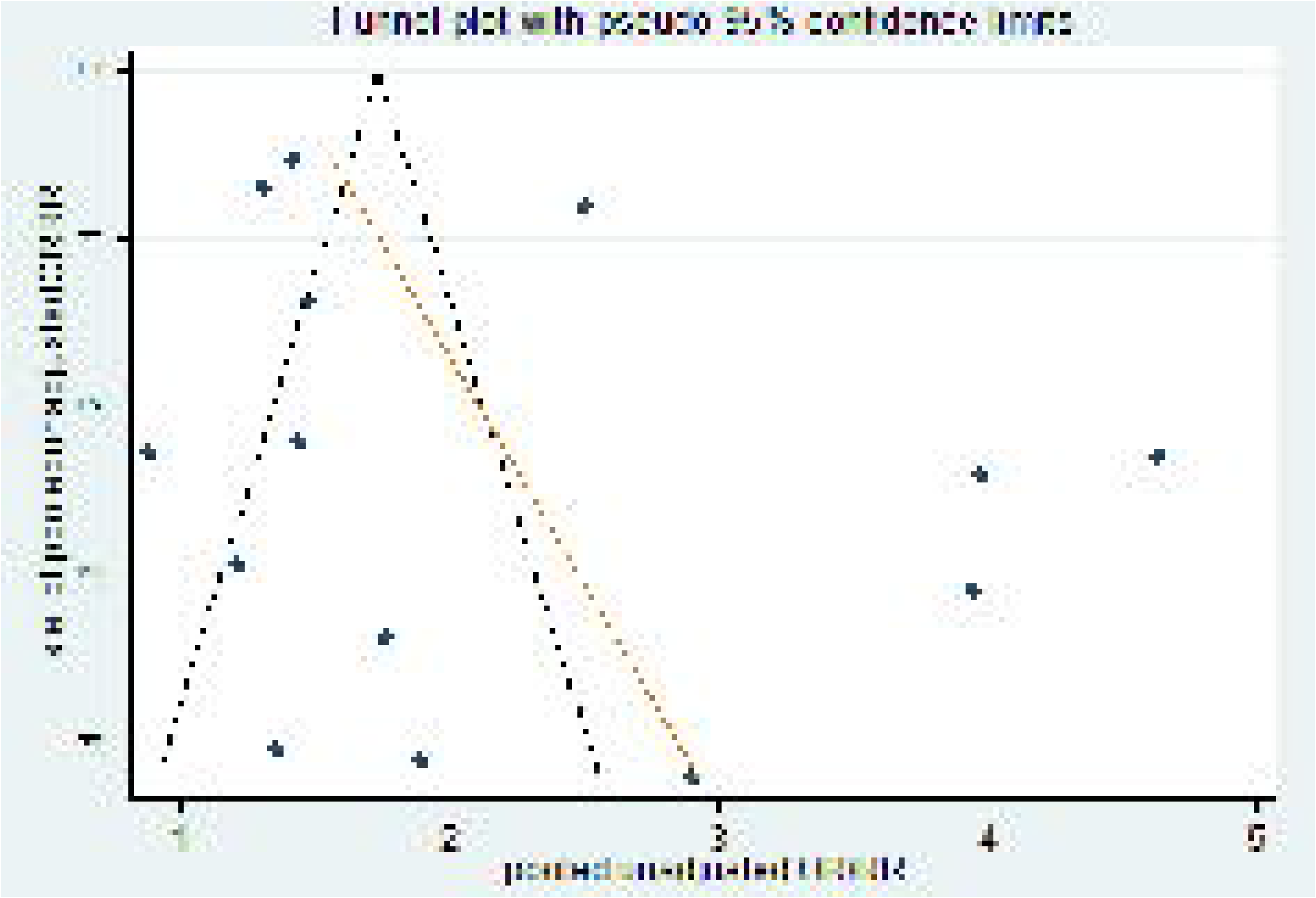
Funnel plot for assessment of publication bias

### Meta-regression analysis

A study level meta-regression analysis was performed for all the important covariates (age: p=0.474, female sex: p=0.766, hypertension: p=0.524, coronary artery disease: p=0.808, heart failure: p=0.263, chronic kidney disease: p=0.875, history of stroke: p=0.252, smoking history: p=0.639, COPD history: p=0.620, and malignancy history: p=0.329). No significant interactions were found between the covariates and the outcome of mortality. The detailed meta-regression results can be found in **Table 3** and **online supplementary material (supplementary figures 1-10)**

**Table 2:** Baseline characteristics of patients per included study.

| Study | Age | Female (n, %) | DM (n, %) | HTN (n, %) | HLD (n, %) | CAD (n, %) | HF (n, %) | CKD (n, %) | CVA (n, %) | Smoking (n, %) | COPD (n, %) | Malignancy (n, %) |
| --- | --- | --- | --- | --- | --- | --- | --- | --- | --- | --- | --- | --- |
| Guan | 48.9±16.3<sup>a</sup> | 674 (42.7) | 130 (8.2) | 269 (16.9) | NA | 59 (3.7) | NA | NA | 30 (1.9) | 111 (7.0) | 24 (1.5) | NA |
| Han | 60 (49-70)<sup>b</sup> | 132 (43.1) | 129 (42.2) | 119 (38.9) | NA | 25 (8.2) | NA | 4 (1.3) | 11 (3.6) | 5 (1.6) | 18 (5.9) | 18 (5.9) |
| Zeng | 67 (57-75)<sup>b</sup> | 38 (39.2) | 26 (26.8) | 47 (48.5) | NA | NA | NA | 8 (8.2) | NA | NA | 8 (8.2) | NA |
| Nikpouraghdam | 55.5±15.2<sup>a</sup> | 1,009 (34.0) | 113 (3.8) | 59 (1.9) | NA | 37(1.3) | NA | 18 (0.6) | NA | NA | 60 (2.0) | 17 (0.6) |
| Javanian | 60.1±13.9<sup>a</sup> | 49 (49.0) | 37 (37.0) | 32 (32.0) | NA | NA | NA | 12 (12.0) | 3 (3.0) | NA | 12 (12.0) | 4 (4.0) |
| Rossi | 63.2<sup>c</sup> | 418 (38.9) | 175 (16.3) | 280 (26.0) | 85 (7.9) | 115 (10.6) | 96 (8.9) | 45 (4.2) | NA | NA | 91 (8.5) | 167 (15.5) |
| Borobia | 61 (46-78)<sup>b</sup> | 1152 (51.8) | 381 (17.1) | 920 (41.3) | NA | NA | NA | 174 (7.8) | NA | 157 (7.1) | 153 (6.9) | 385 (17.3) |
| Perez Guzman | 67 (41-93)<sup>b</sup> | 198 (38.0) | 138 (26.5) | 187 (36.0) | 82 (16.0) | 43 (8.2) | 21 (4.0) | 70 (13.4) | 34 (6.5) | NA | 20 (3.8) | 46 (8.8) |
| Tomlins | 75 (59-82)<sup>b</sup> | 35 (37.0) | 37/95 (38.9) | 35 (37.0) | NA | 21 (22.0) | 15 (16.0) | 22 (23.0) | 8 (8.4) | NA | 10 (11.0) | 20 (21.0) |
| Levy | 21-106<sup>d</sup> | 2,176 (41.6) | 1414 (27.0) | 2474 (50.2) | NA | 454 (9.2) | 219 (4.4) | 326 (6.6) | NA | NA | NA | NA |
| Wang | 60 (46-71) vs. 75 (65-84)<sup>e</sup> | 1,399 (42.7) | 768 (23.5) | 1082 (33.0) | NA | NA | NA | NA | NA | 116 (3.5) | NA | 233 (7.1) |
| Cummings | 62 (51-72)<sup>b</sup> | 87 (34.0) | 92 (35.8) | 162 (63.0) | NA | NA | NA | 37 (14.0) | NA | NA | 24 (9.0) | 18 (7.0) |
| Palaiodimos | 64 (50-73.5)<sup>b</sup> | 102 (51.0) | 79/200 (39.5) | 152 (76.0) | 92 (46.2) | 33 (16.5) | 34 (17.0) | 58 (29.0) | 22 (11.0) | 65 (32.5) | 28 (14.0) | 11 (5.5) |
| Bode | 65 (24-95) vs. 61 (18-101)<sup>f</sup> | 498 (44.4) | 184 (32.3) | NA | NA | NA | NA | NA | NA | NA | NA | NA |

Abbreviations: DM=diabetes mellitus, HTN=hypertension, HLD=hyperlipidemia, CAD=coronary artery disease, HF=heart failure, CKD=chronic kidney disease, CVA=cerebrovascular accident, COPD=chronic obstructive pulmonary, NA=non-available
Notes: a. mean±SD, b. median (IQR), c. mean, d. median (range) in diabetics vs. non-diabetics, d. range, e. median (IQR) in discharged vs. deceased, f. median (range) in diabetics vs. non-diabetics

**Table 3:** Results of the meta-regression analysis.

| **Variables** | **Coefficient** | **Standard error** | **p-value** |
| --- | --- | --- | --- |
| Age | -0.019 | 0.026 | 0.474 |
| Female | -0.003 | 0.010 | 0.766 |
| HTN | -0.005 | 0.007 | 0.524 |
| CAD | 0.594 | 0.361 | 0.808 |
| HF | 0.026 | 0.019 | 0.263 |
| CKD | -0.003 | 0.018 | 0.875 |
| CVA | -0.078 | 0.058 | 0.252 |
| Smoking | -0.012 | 0.024 | 0.639 |
| COPD | -0.022 | 0.043 | 0.620 |
| Malignancy | 0.025 | 0.024 | 0.329 |

Abbreviations: HTN=hypertension, CAD=coronary artery disease, HF=heart failure, CKD=chronic kidney disease, CVA=cerebrovascular accident, COPD=chronic obstructive pulmonary disease

## Discussion

Our study was a systematic review and meta-analysis of observational studies reporting on the association between diabetes and mortality in adult hospitalized patients with COVID-19. Our findings can be summarized as following: i) overall, death was 65% more likely to occur in diabetic inpatients compared to non-diabetic ones but was limited by significant heterogeneity, ii) this association remained significant when the analysis focused on geographical regions of study origin with, once again, significant heterogeneity, iii) our meta-regression analysis did not show an association between how frequent the other significant comorbidities were across different studies and our results.

The findings of our meta-analysis are consistent with the results of some of the large observational cohorts. In one of the largest retrospective studies of hospitalized patients with COVID-19 in New York, diabetics were 33.8% (1,808/5,700) of the total inpatient population (25), whereas the prevalence of diabetes in the general population in New York is 10.5% (26). An early cohort of 1,099 patients with COVID-19 from China revealed that 17.8% of the whole cohort developed severe disease, while the respective rate in the diabetic subgroup was 34.6% (27). One the other hand, another large study from New York (4,103 patients) showed that diabetes is not an independent risk factor for development of critical illness, although a trend was noted (OR: 1.14, 95% CI: 0.83-1.58). (28). Smaller cohorts included in this meta-analysis did not show an association of diabetes with in-hospital mortality, but these studies had relatively small samples and were likely underpowered (18, 22, 23). Given the heterogeneity of the results across the literature, our meta-analysis comes to answer this significant question by utilizing a total sample of 18,506 patients.

Diabetes is associated with higher susceptibility to infectious diseases and higher infection-related mortality (29). A retrospective matched-control study from Canada of more than one million participants showed that diabetics had significantly higher risk to be hospitalized due to an infection, develop sepsis, and die regardless of the affected system or organ or whether the infection was viral or bacterial (30). Similarly, an English cohort of more than 100 thousand diabetics and 200 thousand control subjects revealed that diabetic patients had significantly higher rates of all types of infections with an almost double risk for hospitalization and death compared to non-diabetics (31). Among others, diabetics had 40% higher rates of lower respiratory infections (101.1 vs. 73.3; per 1,000 patients/year) (31). In the 2009 H1N1 pandemic, it was noted that diabetes tripled the risk of hospitalization and quadrupled the risk of admission to intensive care units (32).

Reduced T-lymphocytes response, decreased neutrophil function, impaired humoral immunity, increased adherence of microorganisms in diabetic cells, and increased virulence of some microorganisms in a high-glucose environment are some of the pathogenetic mechanisms that make diabetics more susceptible to infections (4, 33). In addition, diabetes is a major cause of endothelial dysfunction (34). The increasing evidence of endothelial involvement in severe COVID-19 (35, 36), which potentially contributes to COVID-19-associated coagulopathy (37), could raise the hypothesis that the diabetic dysfunctional endothelium is more susceptible to further damage related to COVID-19.

The main strengths of our study are its strict methodology, robust analysis, and the relatively large number of included studies and overall patient sample. Notably three continents and most of the countries that had high COVID-19 incidence were represented. Sensitivity, subgroup, and meta-regression analyses were performed as needed.

The main limitation of our study is the lack of data on glucose control prior or during hospitalization. Therefore, we could not estimate the associations of controlled and uncontrolled diabetes with in-hospital mortality and we recognize that the association would probably be stronger in patients with uncontrolled diabetes and weaker in patients with controlled diabetes. A patient-level meta-analysis would be needed to assess this very important parameter. Second, the estimated association is not adjusted for other important covariates. Unfortunately, only three of the included studies provided adjusted estimates. We tried to solve this methodological issue by performing a meta-regression analysis which showed that the different rates of the other major comorbidities across different studied did not have an impact on the results. Similarly, a patient-level meta-analysis would be the ideal way to adjust for other significant covariates. Third, our meta-analysis was limited by significant heterogeneity, which we tried to assess by using a random-effects model, performing subgroups and sensitivity, and meta-regression analysis.

In conclusion, the present systematic review and meta-analysis revealed that the likelihood of death is 65% higher in diabetic hospitalized patients with COVID-19 compared to non-diabetics. Further studies are needed to assess whether this association is independent or not, as well as to investigate to role of glucose control prior or during the disease. In the meantime, attention should be paid in preventing diabetes and its complications and protecting this population from COVID-19 given the higher chance for adverse outcomes once they are diagnosed with the disease. In addition, diabetic patients diagnosed with COVID-19 should be treated with attention given the possible higher risk for adverse outcomes. While we recognize the limitations, we hope that our study will add to the continuous research of the effect of diabetes in COVID-19.

## Data Availability

Data are available upon request to the first author

## Relationships with Industry and Conflict of Interest

None of the authors

**Online supplementary material**. Ten graphs of the meta-regression analyses for age, female sex, hypertension, coronary artery disease, heart failure, chronic kidney disease, history of stroke, smoking history, COPD history, and malignancy, respectively.

